# The Effect of Intradialytic Exercise on Dialysis Patient Survival: A Randomized Controlled Trial

**DOI:** 10.1101/2022.09.10.22279785

**Authors:** Mohammad Ali Tabibi, Bobby Cheema, Nasrin Salimian, Hugo Corrêa, Saghar Ahmadi

## Abstract

**Introduction:** Patients with kidney failure have a high mortality rate. This study aimed to evaluate the effect of intradialytic exercise on survival in patients receiving hemodialysis (HD).

**Methods:** In this randomized controlled trial conducted in a HD center in Iran, adult patients receiving chronic HD were randomized to intradialytic exercise (60 minutes) in the second hour of thrice weekly dialysis for 6 months (intervention) or no intradialytic exercise (control). The primary outcome was survival rate at 12 months. Secondary outcomes were serum albumin, hemoglobin, hematocrit, red blood cell count, serum calcium, serum phosphorous, parathyroid hormone, physical function (6-minute walk test) and nutritional status (Geriatric Nutritional Risk Index) during the first 6 months. The trial follow-up period was 12 months.

**Results:** The study included 74 participants randomized to intervention (n=37) or control (n=37). Compared with controls, 1-year survival was higher in the intervention group compared with the control group (94% vs 73%, P=0.01). The hazard ratio on univariate analysis in intervention group was 0.17 (95% CI 0.04-0.8; P =0.02) compared to that in control group. During the 6-month intervention period, significant between-group changes were observed in all secondary outcomes between the intervention and control groups.

**Conclusion:** Intradialytic exercise performed for at least 60 minutes during thrice weekly dialysis sessions improves survival in adult patients receiving HD. This effect may be mediated by exercise-related improvements in bone mineral metabolism, anemia, physical function or nutritional status.

**Trial registration:** ClinicalTrials.gov Identifier: NCT04898608, Registered on May 21st,2021. Registered trial name: The Effect of Intradialytic Exercise on Dialysis Patients Survival.

## Introduction

Chronic kidney disease (CKD) is a growing, global public health problem leading to an exponential increase in the number of patients experiencing kidney failure requiring treatment with life-saving kidney replacement therapy (KRT), including hemodialysis (HD), peritoneal dialysis (PD) or kidney transplantation (KT) [1]. HD is the major treatment modality worldwide, accounting for 69% of all KRT and 89% of all dialysis, and is associated with high rates of functional impairment, morbidity, hospitalization, and mortality (10-30 times higher than people with normal kidney function) [2-5]. These outcomes are an urgent priority for patients, caregivers and healthcare professionals [6,7].

A significant factor underpinning these poor outcomes in HD patients is low physical activity related to high rates of comorbidities (such as cardiovascular disease), protein energy wasting, sarcopenia, decreased physical function, decreased aerobic capacity, enforced inactivity during thrice weekly HD sessions, and post-dialysis fatigue [3,8]. Indeed, the activity rates of patients receiving HD is between 20-50% that of healthy people [9]. Prospective observational studies of patients receiving HD have reported a dose-dependent association between physical activity and both all-cause mortality and cardiovascular mortality [9,10]. Increasing physical activity through regular exercise may therefore be an important strategy for improving outcomes in patients receiving HD. Exercise training, particularly a combination of resistance and aerobic exercises, has been reported to improve a number of parameters in HD patients, including physical function, dialysis small solute clearance, mood, appetite, nutrient intake and quality of life [11-13].

A systematic review and meta-analysis of 20 randomized controlled trials involving 677 participants receiving HD demonstrated that exercise training increased exercise capacity (peak VO_2_), walking capacity (6-minute walk test) and both the physical and mental component scores of health-related quality of life (SF-36) [11]. However, the certainty of evidence was substantially reduced by high risks of bias, imprecision and inconsistency (heterogeneity). Moreover, the effects of exercise on mortality in HD patients remain uncertain.

The aim of this study was to evaluate the effects of intradialytic exercise on survival in patients receiving HD. A secondary aim of the study was to assess the effect of intradialytic exercise on surrogate outcomes associated with patient survival.

## Methods

### Trial design

This study was an open-label, parallel arm, randomized controlled trial with blinded end-points, which was conducted in a medical center in Iran. The study protocol was approved by Iran National Committee for Ethics in Biomedical Research (approval number IR.IAU.KHUISF.REC.1399.146) and was conducted in accordance with principles of the Declaration of Helsinki. Recruitment occurred between January 25, 2020 and 2 February 2020. Follow-up continued until August 5, 2021.

### Participants

Individuals were eligible to participate in the study after meeting all of the following inclusion criteria: 1) age ≥ 18 years; 2) receiving regular HD 3 times a week; 3) on HD for at least 1 year, 4) absence of a history of myocardial infarction within the past 3 months; 5) permission from their doctors to participate; and, 6) had capacity to provide informed consent to participate in the study. Individuals were excluded if they met any of the following exclusion criteria: 1) cardiac instability (angina, decompensated congestive heart failure, severe arteriovenous stenosis, uncontrolled arrhythmias, etc.); 2) active infection or acute medical illness; 3) hemodynamic instability; 4) labile glycemic control; 5) inability to exercise (e.g. lower extremity amputation with no prosthesis); 6) severe musculoskeletal pain at rest or with minimal activity; 7) inability to sit, stand or walk unassisted (walking device such as cane or walker allowed); or, 8) shortness of breath at rest or with activities of daily living (NYHA Class IV).

### Trial procedures

After providing written informed consent, eligible patients received a baseline assessment. Data were collected on demographic characteristics (age, sex, and time on hemodialysis), primary cause of kidney failure, and comorbidities (atherosclerotic heart disease, congestive heart failure, cerebrovascular accident/transient ischemic attack, peripheral vascular disease, dysrhythmia, and other cardiac diseases, chronic obstructive pulmonary disease, gastrointestinal bleeding, liver disease, cancer, and diabetes). Comorbidities were quantified using Charlson comorbidity index (CCI) established for dialysis patients, which included the underlying cause of kidney failure, as well as 11 comorbidities [14].

Participants were then randomized in a 1:1 ratio to either the intervention group or control group. The randomization sequence was generated by a study biostatistician who was not otherwise involved in the study using a computer-generated random schedule (using Stata 16, Stata Crop, College Station, Tx). Allocation concealment was safeguarded through the use of sequentially numbered, sealed, opaque envelopes by a specified staff member who was not involved in the study..

### Intervention

Subjects in the intervention group performed concurrent intradialytic exercise during the 2nd hour of dialysis (60-minute exercise sessions three times a week) for 6 months. The intervention was a combination of aerobic and resistance exercises. Workout time at the beginning was 30 minutes and gradually increased to 60 minutes. Exercises were individualized in a way that matched the level of physical fitness of participants (Supplementary Appendix). Aerobic exercises consisted of continuously performed specified movements, such as moving legs back and forth, shoulder abduction and adduction (hand without fistula), flexing and extending the knee, internally and externally rotating the leg, and abducting and adducting the leg, in time with a played beat.

The rhythm of continuous movements was adjusted by the beats per minute of the music. This meant that participants had to coordinate the movements of their arms and legs with the beats per minute of the song being played to them. In this way, the speed and intensity of aerobic exercise was controlled by the rhythm. Resistance training was performed in a semi-recumbent position and included exercises for the upper and lower limbs as well as core strength exercises using body weight, weight cuffs, dumbbells, and elastic bands of varying intensity. Leg abduction, plantarflexion, dorsiflexion, straight-leg/bent knee raises, knee extension, and knee flexion were all part of the resistance training program.

Participants in the control group did not undertake any specific physical activity during dialysis. All participants were followed for 12 months.

All other pharmacological, dialysis, dietary and management protocols were identical for participants in both groups. All participants received normal bicarbonate hemodialysis, which was carried out three times a week for an average of 4 hours. Volumetric ultrafiltration control was available on all machines. The standard dialysate flow rate was 500 mL/min and blood flow rates were prescribed according to the participant’s needs. Automated methods were applied to perform dialyzer reuse uniformly.

### Blood sampling

Baseline blood samples were collected one day before the start of the exercise session the day before a mid-week session. Exercise began at the mid-week dialysis session. After the end of the 36th session (end of the third month) and after the end of the 72nd session (end of the sixth month), subsequent blood samples were collected before the midweek dialysis session. The control group was assessed at the same time points. On a nondialysis day, blood samples were taken from the arterial needle after at least an 8-hour fast. Approximately 30 milliliters of blood were collected and centrifuged for 15 minutes at 20 °C and 2500 g. Plasma was next pipetted into cryotubes and stored at -80 °C in a freezer that was electronically monitored. All samples were measured in duplicate, in line with the manufacturers’ suggested protocol, and within the manufacturer’s specified range of acceptable variation and sensitivity.

### Outcomes

The primary outcome measure was 1-year survival. Time frame started by ending the intervention (6th month). Information about the time and cause of death were extracted from participants’ medical records. This included information recorded in the CMS-2746 form.

Secondary outcome measures included changes in serum albumin (g/dL), hemoglobin (g/dL), hematocrit (%), red blood cell count (×10^6^/μL), serum calcium (mg/dL), serum phosphorous (mEq/L), and parathyroid hormone (PTH) (pg/mL) over time. Other secondary outcomes were physical function and nutritional status. These outcomes were evaluated at baseline, 3 months and 6 months.

Physical function was evaluated with the 6-minute walk test (6MWT), which is a functional examination of exercise capacity and was performed according to the American Thoracic Society guidelines [15]. In brief, the test was performed indoors on a 30m straight course. Participants were instructed to walk as fast as possible for 6 minutes. Walking aids were allowed and recorded. At the end of the 6-minute period, the distance was measured.

Nutritional status was assessed by Geriatric Nutritional Risk Index (GNRI), as reported by Bouillanne et al. [16] and modified for older patients, as reported by Yamada et al. [17]. The index was calculated as follows: GNRI = (14.89 albumin (g/dlL) + (41.7 x [body weight/ideal body weight]). When a participant’s actual weight was greater than their ideal weight, the body weight/ideal body weight ratio was set to 1 by default. Instead of utilizing the Lorentz formula from the original GNRI equation to determine the ideal body weight, the value obtained from the participant’s height and a body mass index of 22 was used [16]. Lower GNRI is a significant predictor of bone mineral disorders, cardiovascular events and all-cause mortality in dialysis patients [18-20].

Safety outcomes included all serious adverse events and adverse events.

### Blinding

Due to the nature of the intervention, it was not feasible to blind participants or study staff. However, outcome assessors and data analysts were blinded to participants’ treatment allocations.

### Sample size

The sample size was calculated by NCSS PASS 16.0 software. The model was established according to the log-rank test of survival analysis (bilateral side), with α=0.05 and power 1 – β = 0.8. Due to the fact that there had been no prior randomized controlled trials evaluating the effect of exercise on survival in dialysis patients, the results of a non-randomized study of the association of changes in physical activity with survival in dialysis patients were used to inform sample size calculations [21]. Assuming a hazard ratio of 0.25 between the intervention and control groups and a drop-out rate of 20%, 74 participants (37 per group) were required to provide 80% power.

### Statistical analysis

Data are presented as frequency (percentage), mean ± standard deviation, or median and interquartile range, depending on data type and distribution. A detailed statistical analysis plan was prepared and completed prior to database lock. The primary outcome of survival was analyzed by Kaplan-Meier analysis and log rank test. Overall survival was calculated from the date of end of intervention to the date of death from any cause. Participants remaining alive were censored at the date of last follow-up. Cox regression model was constructed to evaluate the effect of intervention on survival rate. Secondary outcomes were evaluated using Repeated Measure ANOVA and the Friedman test. Statistical analyses were performed using IBM SPSS software 25. P values less than 0.05 were considered statistically significant.

## Results

Overall, 95 patients were assessed for eligibility, of whom 74 were consented and randomized. The corresponding flowchart is presented in Figure 1. During the 6-month intervention period, 2 participants in the intervention group and 4 participants in the control group were excluded from the intervention. Every patient who was lost to follow-up (for any reason except death) during 12-month follow-up was censored.

**Figure 1.**
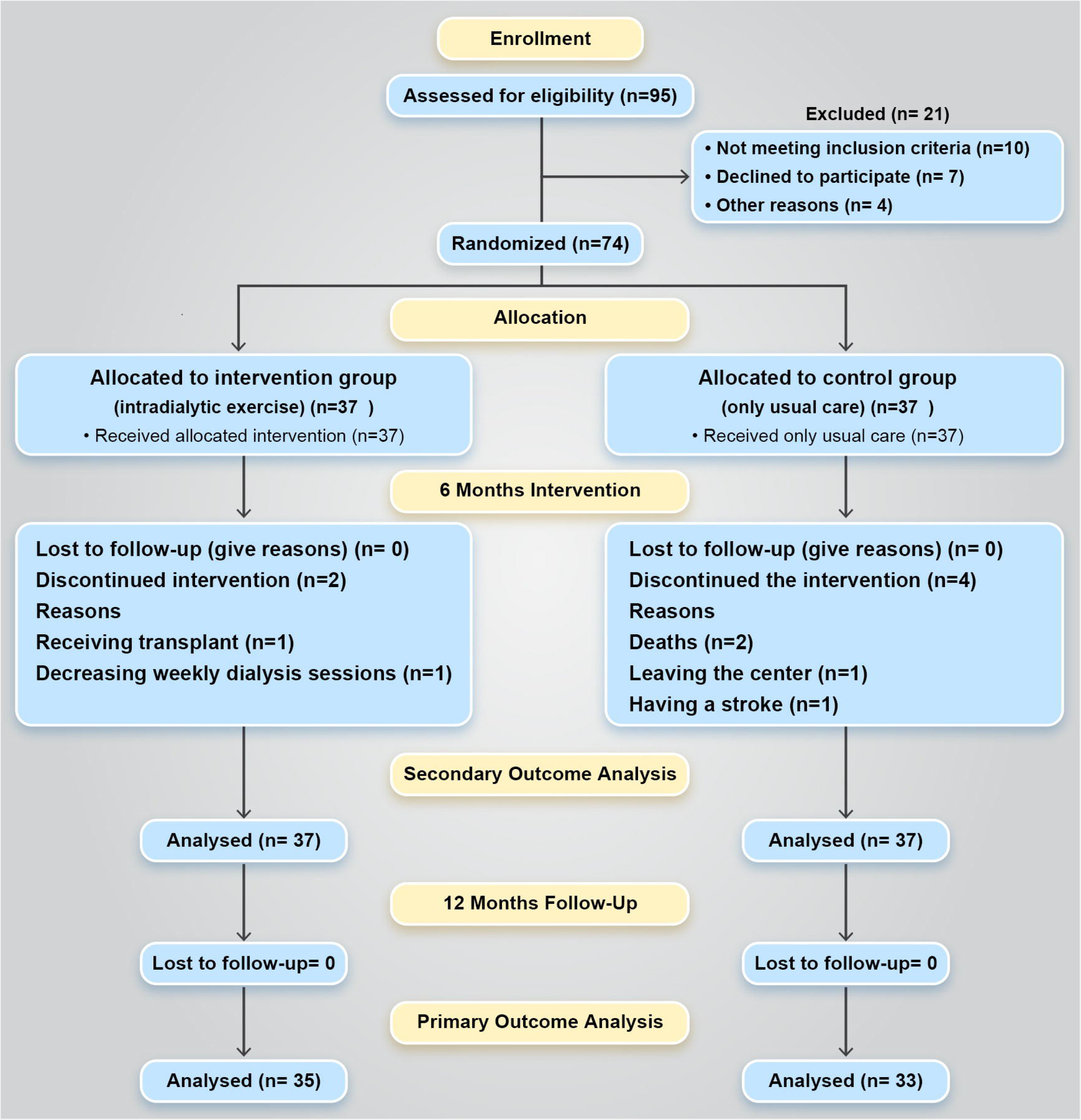
Participant flow during the study.

## Baseline characteristics

Baseline characteristics were balanced between the assigned treatment groups (Table 1.)

**Table 1.** Baseline characteristics of patients

|  | Intervention (n=37) | Usual Care (n=37) |
| --- | --- | --- |
| Sex Male | 21 (57%) | 23 (62%) |
| Age (year) | 62 ± 13 | 65 ± 11 |
| Hemodialysis history (months) | 29 ± 10 | 26 ± 9 |
| Primary kidney disease |  |  |
| Diabetes | 16 (43%) | 20 (54%) |
| Hypertension | 10 (27%) | 11 (30%) |
| Glomerulonephritis | 5 (14%) | 3 (8%) |
| Other | 6 (16%) | 3 (8%) |
| CCI | 6 ± 2 | 7 ± 2 |

Values are as mean (standard deviation) or n (%)
CCI Charlson Comorbidity Index

### Primary outcome

Before the end of initial 6-month period during which the exercise intervention was being performed, no one in the intervention group and two participants in the control group died.

Two participants (6%) in the intervention group died during the follow-up period due to cardiovascular disease (n=1) and unknown cause (n=1). In contrast, 9 participants in the control group died due to cardiovascular disease (n=3), cerebrovascular disease (n=2), infection (n=1), other causes (n=2) and unknown cause (n=1). The cumulative survival rate in the control group was significantly lower than that in the intervention group (Log rank statistics = 6.5, P=0.01) (Fig 2). Similar results were found in univariable Cox regression model analyses (Table 2). The hazard ratio (HR) for mortality in the intervention group was 0.17 (95% confidence interval [CI] 0.04-0.8, P =0.02), compared to the control group.

**Figure 2.**
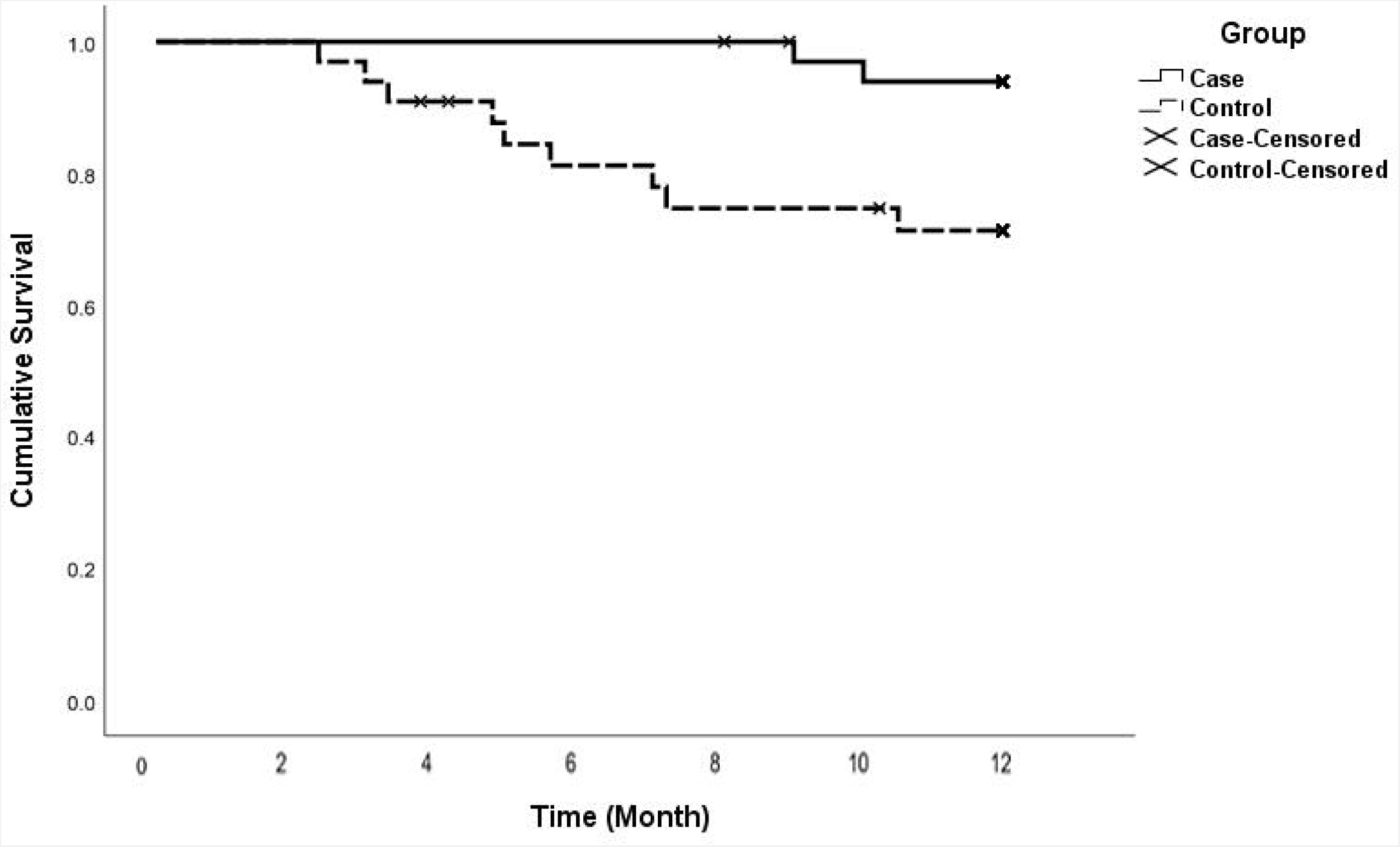
Kaplan-Meier survival curve for patients randomly allocated to intradialytic exercise for 6 months (intervention) or usual care (control). The difference between the groups was statistically significant (p=0.01).

**Table 2.** Cox regression model for all-cause mortality

| Parameter | Univariate analysis |  |  |
| --- | --- | --- | --- |
|  | Hazard ratio | (95% CI) | P value |
| Intervention group | 0.17 | (0.04-0.8) | 0.02* |
| Control group | 1.00 |  |  |

\*p < 0.05 significant

### Secondary outcomes

Significant between-group changes during the 6-month intervention period were observed in all secondary outcomes (Table 3) between the intervention and control groups. Specifically, serum albumin, hemoglobin, red blood cell count, serum calcium, physical function (6MWT) and nutritional status (GNRI) tended to increase in the intervention group, but remained relatively stable in the control group. In contrast, serum parathyroid hormone and phosphorus levels significantly decreased in the intervention group but remained relatively stable in the control group (Table 3).

**Table 3.** Change of secondary outcomes and the results of longitudinal analysis

|  | Intervention |  |  | Usual care |  |  | Group<br>×Time |
| --- | --- | --- | --- | --- | --- | --- | --- |
|  | Baseline<br>(n=37) | 3 months<br>(n=36) | 6 months<br>(n=35) | Baseline<br>(n=37) | 3 months<br>(n=35) | 6 months<br>(n=33) |  |
| Alb (g/dl) | 3.5 ± 0.3 | 3.7 ± 0.3 | 4 ± 0.3 | 3.7 ± 0.3 | 3.6 ± 0.3 | 3.5 ± 0.2 | <0.001** |
| HGB (g/dl) | 10 ± 1 | 10.8 ± 1.3 | 11.7 ± 1.4 | 10.8 ± 1.3 | 10.2 ± 1.2 | 9.9 ± 1.2 | <0.001** |
| RBC<br>(10<sup>6</sup>/μL) | 3.5 ± 0.4 | 3.9 ± 0.5 | 4.2 ± 0.5 | 3.8 ± 0.5 | 3.6 ± 0.5 | 3.5 ± 0.5 | <0.001** |
| HCT (%) | 31.8 ± 3.2 | 34.3 ± 3.4 | 36.8 ± 3.7 | 33.9 ± 3.2 | 32.2 ± 3.1 | 31.1 ± 3 | <0.001** |
| Ca (mg/dl) | 7.9 ± 0.9 | 8.6 ± 7 | 9.1 ± 0.6 | 8.6 ± 0.8 | 8.6 ± 0.7 | 8.3 ± 0.7 | <0.001** |
| P (meq/lit) | 5.6 ± 1.5 | 5 ± 1.2 | 4.3 ± 0.6 | 5.4 ± 1.4 | 5.9 ± 1 | 5.3 ± 1 | <0.001** |
| PTH (pg/ml) | 441<br>(264 - 567) | 290<br>(224 - 391) | 245 †<br>(173 - 286) | 270<br>(180 - 418) | 342<br>(245 - 476) | 312<br>(243 - 407) | NA |
| GNRI | 92.8 ± 4.6 | 96.1 ± 5.1 | 101.1 ± 4.1 | 96 ± 5.5 | 94.5 ± 4.9 | 92.7 ± 5 | <0.001** |
| 6 mwt (m) | 309 ± 86 | 348 ± 92 | 381 ± 99 | 323 ± 78 | 304 ± 83 | 283 ± 87 | <0.001** |

Alb albumin, HGB hemoglobin, RBC Red blood cells, HCT hematocrit, Ca serum calcium, P serum phosphorous, PTH parathyroid hormone, GNRI Geriatric Nutrition Risk Index, 6 mwt 6-min walk test
Values are as mean ± standard deviation or median (interquartile)
Group ×Time is an interaction from a two-way repeated-measures analysis of the effect of time from baseline to 6 months
\*p < 0.05 significant
\*\*p < 0.01 highly significant
†p < 0.01 highly significant within-group difference, analyzed by Friedman test

### Safety

No treatment-related serious adverse effects were observed during the period of the study. During the intervention period only one of the patients had muscle cramp after two sessions of the exercise and one of the patients had bleeding from fistula when she was doing the exercises. Of course, it was not serious or harmful.

## Discussion

This study showed that intradialytic exercise for 6 months improved subsequent survival in adult patients receiving HD for 12 months. Furthermore, compared with controls, intradialytic exercise caused potentially beneficial improvements in selected laboratory parameters (serum albumin, hemoglobin, red blood cell count, serum calcium, serum PTH, serum phosphorus), physical function (6MWT) and nutritional status (GNRI) during the 6-month intervention period compared with controls.

The finding of a survival benefit from intradialytic exercise is in keeping that of with previous observational studies investigating the association between increased physical activity and survival in patients receiving dialysis [10,22]. In a prospective, multinational study of 8043 patients receiving HD in centers across Argentina and Europe, Bernier-Jean et al [10] found that, compared with self-reported physical inactivity, occasional activity (≤1x/week) and frequent activity (>1x/week) were dose-dependently associated with lower all-cause mortality and cardiovascular mortality, but not non-cardiovascular mortality. This study was potentially limited by social desirability bias, misclassification bias and residual confounding from unmeasured determinants of better health. Similarly, in a systematic review of 11 observational studies involving patients with kidney failure (6 in HD patients, 3 in kidney transplant recipients, 2 in both HD and PD patients), Martins et al [22] demonstrated that physical activity (self-reported in 10 studies) was generally inversely associated with mortality. However, high degrees of statistical, clinical and methodological heterogeneity precluded meta-analysis. This fact, together with the observational designs of the included studies meant that the evidence was very low certainty. Recently, Mallalamaci et al [23] reported long-term (36 month) follow-up outcomes of the EXCITE study, a prospective, randomized controlled trial of a 6-month home-based walking exercise program in 227 patients on dialysis (192 on HD) at 13 Italian centers. Compared with controls, patients allocated to the exercise arm had a borderline significantly lower risk of the composite outcome of hospitalization or death (HR 0.71, 95% CI 0.50 – 1.00). As this was a secondary outcome of the trial, the findings were hypothesis-generating only. Until the present study, the effect of intradialytic exercise on patient survival had not been evaluated by randomized controlled trial.

The mechanisms underpinning the improved survival of patients performing intradialytic exercise for 6 months may be explained by improvements in a number of factors associated with physical exercise including bone mineral metabolism [24-27], anemia [28], cholesterol [29], endothelial function (by raising the laminar sheer stress bioavailability of nitric oxide) [30], reduced arterial stiffness due to nitric oxide-mediated vasodilation [29], improved coronary blood flow reserve [29], reduced blood pressure [29], increased physical function [31], reduced oxidative stress [29], decreased inflammation [32], reduced sympathetic nervous system activity [29], and promotion of antithrombotic actions that favor fibrinolysis over thrombosis [30]. In the present study, significant improvements were observed in bone mineral metabolism, anemia, nutrition, physical function (as measured by the 6MWT) and nutritional status (as measured by GNRI). Each of these surrogate outcome measures have been strongly associated with survival in patients receiving HD [33-42].

A major strength of the present study was the fact that all exercises were tailored according to each individual’s functional status within a pre-specified structure. There was also a high participation rate, such that included HD patients exhibited considerable diversity with respect to demographic characteristics and associated comorbidities. The study also evaluated a range of biochemical and functional parameters to explore the potential mechanisms of any effect of intradialytic exercise on patient survival. The intervention was designed so that it was broadly implementable in most HD centers, including in low resource settings.

Balanced against these strengths, the study also had a number of limitations. The duration of the intradialytic exercise intervention was short (6 months), as was the subsequent follow-up (12 months), meaning that the long-term effects of exercise on patient survival remain uncertain. The small sample size led to an imprecise estimate of the effect of exercise on patient survival.

Survival was timed from the end of the intervention, which introduced immortal time bias. Secondary outcome measures exploring the effects of intradialytic exercise and patient survival were only examined during the first 6 months of the study.

## Conclusion

Intradialytic exercise performed for at least 60 minutes during thrice weekly dialysis sessions improves survival in adult patients receiving HD. This effect may be mediated by exercise-related improvements in bone mineral metabolism, anemia, physical function or nutritional status. Further large-scale studies are warranted.

## Supporting information

CONSORT Checklist

## Data Availability

All data produced in the present study are available upon reasonable request to the authors.

## Acknowledgement

The authors would like to express their heart-felt gratitude to all the investigators for their contribution to the trial, especially Dr. Paul Bennett, Dr. Ken Wilund and Dr. Tomas Storer, also Dr. David W Johnson for critically reviewing and editing the manuscript, the statistical support and all the staff of the dialysis center for their efforts and patience in helping to maintain the standards of the research, as well as all patients involved in this study. Also, the authors would like to thank Pardis Specialized Wellness Institute and all the personnel of the institute for their support throughout the study. The authors would also like to thank the opportunity that medRxiv offered them to deposit the preprint version.

This study is endorsed by the Global Renal Exercise (GREX) network. The interpretation and conclusions contained herein are those of the researchers and do not represent the views of GREX.

## Authors Contribution

MAT: conceptualizing the study, project leader of the study, conducting the study, writing of the manuscript, supervising of the analysis, interpreting the results, approval of the manuscript, BC: conceptualizing of the study, supervision of the study, co-writing of the manuscript, supervising of the manuscript, approval of the manuscript, NS: project leader of the study, analyzing the data, interpreting the results, co-writing of the manuscript, approval of the manuscript, HC: conceptualizing the study, supervising of the study, co-writing of the manuscript, supervising of the manuscript, approval of the manuscript, SA: conceptualizing the study, conducting the study, co-writing of the manuscript, approval of the manuscript. Each author contributed with important intellectual content during the manuscript drafting or revision and accepts accountability for the overall work by ensuring that questions pertaining to the accuracy or integrity of any portion of the work are appropriately investigated and resolved. All authors read and approved the final manuscript.

## Availability of data

The datasets (without any identifying information) used are available from the corresponding author on reasonable request.

## Supplemental Material

### TABLE OF CONTENTS

Supplemental Material: Intervention Protocol

Supplemental Table 1

Supplemental Table 2

Supplemental Figure 1

