## Supplemental file for "The Effect of Intradialytic Exercise on Dialysis Patient Survival: A Randomized Controlled Trial"

**TABLE OF CONTENTS**

Supplemental Material: Intervention Protocol

Supplemental Table 1

Supplemental Table 2

Supplemental Figure 1

**Supplemental Material: Intervention Protocol**

The intervention was a combination of aerobic and resistance exercises during dialysis. All exercises were performed in the second hour of dialysis. Workout duration at the beginning was as long as 30 minutes and gradually increased to 60 minutes. Exercises were individualized in a way that matched the level of physical fitness of patients. Participants allocated to the intervention group performed 6 Minute Walk Test (6MWT), Timed Up and Go (TUG) and 30s Sit to Stand (30sSTS) tests [1]. The range of scores obtained by patients in each test were recorded.

Participants who were in the first tertile in two or three tests, were in the “very low” group, participants who were in the second tertile in two or three tests, were in the “low” group and participants who were in the third tertile in two or three tests were in the “moderate” group. Exercises were further individualized according to cardiovascular risk factors[2,3]. More details are in Supplementary Table 1. If a patient obtained one or more ratios of very high risk or three or more ratios of high risk, the patient’s fitness level was reduced by one category to mitigate the risk of cardiovascular events. The intensities of training for each group and the rates of increases in training durations are presented in Supplementary Figure 1. Exercise training was tailored so that it was perceived by patients as moderately strenuous (Borg Scale of perceived exertion: 12–13, “somewhat hard”).

Each workout session included 5 minutes of warm-up, aerobic exercises, resistance exercises and finally 10 minutes of stretching exercises to cool down.

**Aerobic Exercise**

Aerobic exercises in the form of repetitive specified movements, such as moving legs back and forth, raising and lowering the hand (hand without fistula), bending and straightening the knee, rotating the leg, moving the leg sideways, and the like, were performed continuously in time with a played beat. The rhythm of continuous movement was adjusted according to the frequency (beats per minute) of music, such that patients had to coordinate the movements of their arms and legs with the beat of the played music. Before the intervention, for each group, the specific rhythm of that group was adjusted in such a way that patients could reach their target heart rate by performing movements according to that rhythm. At the end of the third month, each group's specific songs were set again according to progress of the group. The target heart rate (THR) was determined by the Karvonen formula (THR = (220 - Age)*TI). The aerobic training intensities (TI) were adjusted to maintain heart rates between 50% and 60% of maximum heart rate in the very low intensity group, between 60% and 65% of maximum heart rate in the low intensity group, and between 65% and 70% of maximum heart rate in the moderate group. Aerobic exercise started with five minutes per session and gradually increased each month until reaching 15 minutes per session by the third month; thereafter, aerobic exercise session durations were 15 minutes until the sixth month.

**Resistance training**

Resistance training was comprised of exercises for upper and lower limbs performed with body weight, weight cuffs, dumbbells and elastic bands of varying resistance, as well as core strength exercises, in a semi-recumbent position. The resistance training program included: leg abduction, plantar flexion, dorsi flexion, straight-leg/bent knee raise, knee extension and knee flexion. For each patient, the exercises were 60% of 1 repetitio maximum (1RM). For calculating 1RM, related equations were used to predict the 1RM from a multiple-RM test [4]. At the beginning of the program, each type of exercise was performed in 1 set for the very low group, 2 sets for the low group, and 3 sets for the moderate group (Supplemental Figure 1). Given that the total volume of training (resistance and aerobic combination) at the beginning of the program was 30 minutes, the number of repetitions and the number of sets or number of exercises increased gradually every 4 weeks until reaching a maximum by the end of the third month. Supplemental Table 2 and Supplemental Figure 1 provide further details. At the end of the third month, the functional tests were repeated to re-evaluate the patient's condition, and to reassign exercise intensity grouping as needed. 1RM were determined again for each patient. Under these conditions, the weight of the dumbbells and the degree of tension of the elastic bands were increased progressively, which led to an increase in the intensity of the exercise, provided that the patient had been adapted to the previous exercises, and now had the capacity for higher intensity. For participants in the moderate group, at the beginning of 4th month, two new exercises were added. Overall, the perceived exertion was kept the same for all individuals between 12 and 14 on the Borg scale. At the end of each session, the exercise physiologist reviewed the adherence checklists. If a person did attend an exercise session, a counseling session was held with both the nephrologist and exercise physiologist. The reason for the individual's non-participation was investigated and patients were encouraged to continue with their exercise program.

**Supplement Table 1.** Classification by health markers

| Risk Level | Health Markers | | | | | | |
|---|---|---|---|---|---|---|---|
| | BMI | WHR | | WC (cm) | | WHtR | |
| | | Men | Women | Men | Women | Men | Women |
| **Low** | ≤18.49 | ≤0.90 | ≤0.76 | ≤94 | ≤80 | ≤0.49 | ≤0.50 |
| **Medium** | 18.50-24.99 | 0.91-0.93 | 0.77-0.84 | | | 0.50-0.56 | 0.51-0.57 |
| **High** | 25-34.99 | 0.94-1.03 | 0.85-0.9 | 94.1-102 | 80.1-88 | 0.57-0.63 | 0.58-0.65 |
| **Very High** | ≥35.00 | ≥1.04 | ≥1.00 | ≥102.1 | ≥88.1 | ≥0.63 | ≥0.65 |

BMI: body mass index, WC: waist circumference, WHR: Waist-to-Hip Ratio, WHtR: Waist-to-height Ratio, cm: centimeters.

**Supplemental Table 2.** Intervention design for resistance training

| | Group | | |
|---|---|---|---|
| | **Very low** | **Low** | **Moderate** |
| **Week** | Set*Rep | Set*Rep | Set*Rep |
| **1** | 1*8 | 2*8 | 3*8 |
| **2** | 1*8 | 2*8 | 3*8 |
| **3** | 1*10 | 2*10 | 3*8 |
| **4** | 1*12 | 2*10 | 3*8 |
| **5** | 2*8 | 2*12 | 3*10 |
| **6** | 2*8 | 2*12 | 3*10 |
| **7** | 2*10 | 3*8 | 3*10 |
| **8** | 2*12 | 3*8 | 3*10 |
| **9** | 3*8 | 3*10 | 3*12 |
| **10** | 3*8 | 3*10 | 3*12 |
| **11** | 3*10 | 3*12 | 3*12 |
| **12** | 3*12 | 3*12 | 3*12 |
| | **Performing functional tests** | | |
| **13** | 1*8 | 2*8 | 3*8 |
| **14** | 1*10 | 2*8 | 3*8 |
| **15** | 1*12 | 2*10 | 3*8 |
| **16** | 1*12 | 2*10 | 3*8 |
| **17** | 2*8 | 2*12 | 3*10 |
| **18** | 2*10 | 2*12 | 3*10 |
| **19** | 2*12 | 3*8 | 3*10 |
| **20** | 2*12 | 3*8 | 3*10 |
| **21** | 3*8 | 3*10 | 3*12 |
| **22** | 3*10 | 3*10 | 3*12 |
| **23** | 3*12 | 3*12 | 3*12 |
| **24** | 3*12 | 3*12 | 3*12 |

Rep: Repetitions

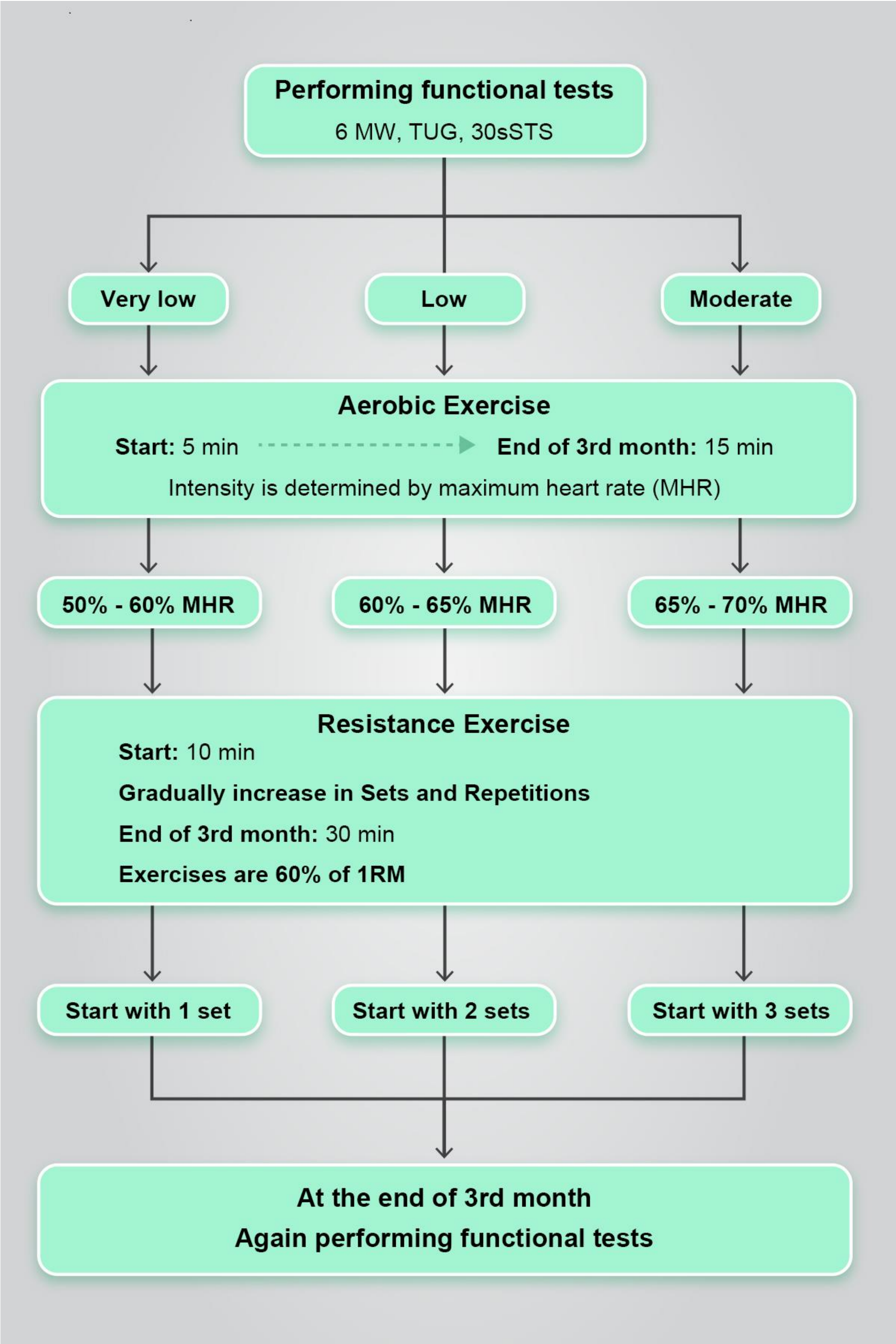


**Supplemental Figure 1.** Intervention design.

6MWT: 6 Minute Walk Test, TUG: Timed Up & Go, 30sSTS: 30s Sit To Stand test, MHR: Maximum Heart Rate, 1RM: one- Repetition Maximum
